# Spatial Growth Rate of Emerging SARS-CoV-2 Lineages in England, September 2020–December 2021

**DOI:** 10.1101/2022.02.02.22270110

**Authors:** M.R. Smallman-Raynor, A.D. Cliff, The COVID-19 Genomics UK (COG-UK) Consortium

## Abstract

This paper uses a robust method of spatial epidemiological analysis to assess the spatial growth rate of multiple lineages of SARS-CoV-2 in the local authority areas of England, September 2020–December 2021. Using the genomic surveillance records of the COVID-19 Genomics UK (COG-UK) Consortium, the analysis identifies a substantial (7.6-fold) difference in the average rate of spatial growth of 37 sample lineages, from the slowest (Delta AY.4.3) to the fastest (Omicron BA.1). Spatial growth of the Omicron (B.1.1.529 and BA) variant was found to be 2.81× faster than the Delta (B.1.617.2 and AY) variant and 3.76× faster than the Alpha (B.1.1.7 and Q) variant. In addition to AY.4.2 (a designated variant under investigation, VUI-21OCT-01), three Delta sublineages (AY.43, AY.98 and AY.120) were found to display a statistically faster rate of spatial growth than the parent lineage and would seem to merit further investigation. We suggest that the monitoring of spatial growth rates is a potentially valuable adjunct to routine assessments of the growth of emerging SARS-CoV-2 lineages in a defined population.

## Introduction

Emerging lineages of the severe acute respiratory syndrome coronavirus 2 (SARS-CoV-2) have the potential to place significant pressure on public health systems due to increased infectivity, transmissibility, virulence, immune escape or other fitness advantage [1, 2]. Global genomic surveillance has identified >1,700 SARS-CoV-2 lineages since the beginning of the COVID-19 pandemic [3, 4], of which Alpha (B.1.1.7 and Q), Beta (B.1.351), Gamma (P.1), Delta (B.1.617.2 and AY) and Omicron (B.1.1.529 and BA) have been designated as variants of concern by the World Health Organization (WHO) on account of their global public health significance [5]. Additional lineages are currently classified on the basis of properties that are suggestive of an emerging (variants of interest) or possible future (variants under monitoring) risk to global public health [5]. The risk is well illustrated by the recent and rapid emergence of Omicron as the dominant variant in the United Kingdom, South Africa and the United States, among other countries, in late November and December 2021 [6–8].

One important epidemiological facet of an emerging SARS-CoV-2 lineage is its propensity to grow in a defined population [9]. There are well-established methods for assessing the rate of temporal growth by, for example, examining the trajectory of case doubling times or estimating the basic reproduction number, *R*_0_, of the agent in question [10, 11]. Viewed from a geographical perspective, these measures are essentially aspatial in that they provide very little information on the geographical growth, or spatial expansion, of the associated infection wave. To extend the examination of SARS-CoV-2 growth rates into the spatial domain, the present paper applies a robust method of spatial epidemiological analysis that is known as the *swash-backwash model of the single epidemic wave* [12] to the genomic surveillance records of the COVID-19 Genomics UK (COG-UK) Consortium [13]. Using the spatial sequence of detection of sample variants as a proxy for the spatial wave front of infection, our examination yields estimates of the spatial growth rate of multiple SARS-CoV-2 lineages in the local authority areas of England, September 2020–December 2021.

For a total of 37 sample lineages under investigation, we present evidence of a substantial (7.6-fold) difference in the average rate of spatial growth, from the slowest (Delta AY.4.3) to the fastest (Omicron BA.1). Whilst the overall results for the Alpha, Delta and Omicron variants are consistent with the documented growth advantages for these lineages, several emergent Delta sublineages (AY.4.2, AY.43, AY.98 and AY.120) are found to have had a statistically significant growth advantage over the parent lineage. To our knowledge, this is the first comparative study of the spatial growth rate of multiple emerging SARS-CoV-2 lineages at the national level. It is also the first report of a spatial growth advantage for the Delta AY.43, AY.98 and AY.120 lineages, and the first to document an apparently reduced spatial growth rate for a substantial number of other AY lineages that emerged in the spring and summer of 2021. We suggest that the monitoring of spatial growth rates is a potentially valuable adjunct to routine assessments of the growth of SARS-CoV-2 lineages in a defined population.

## Data and methods

Since September 2020, successive waves of SARS-CoV-2 infection with emerging lineages of the Alpha (September 2020 onset), Delta (March 2021 onset) and Omicron (November 2021 onset) variants have been recorded in England [14–16]. The weekly record of COVID-19 cases to mid-December 2021 is plotted in Figure 1A, whilst the underpinning sequence of variants is depicted in Figure 1B. As Figure 1B shows, Alpha, Delta and Omicron achieved the status of dominant variants in December 2020, May 2021 and December 2021, respectively.

**Figure 1.**
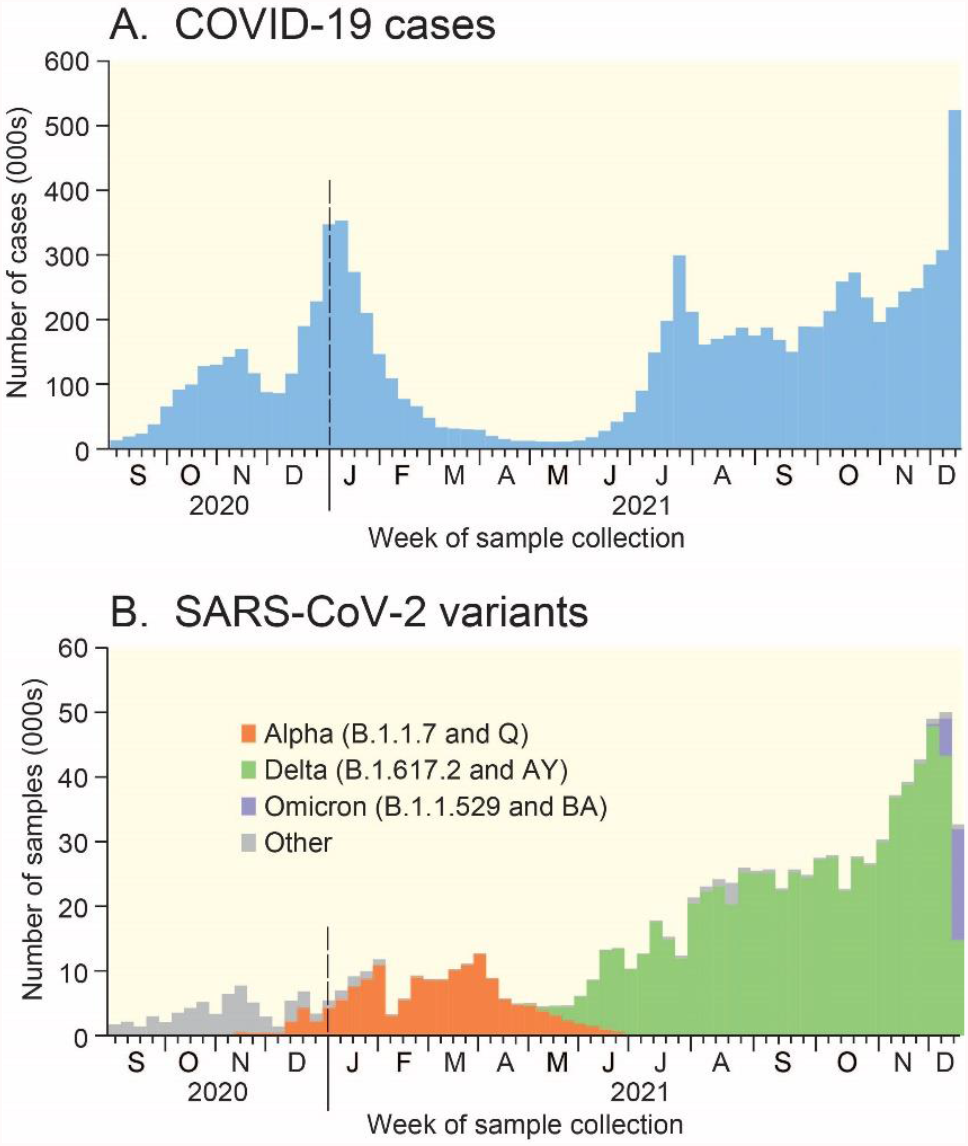
COVID-19 cases in England, September 2020–December 2021. (A) Positive COVID-19 test specimens as recorded by the UK Government. (B) Number of sample genomes of SARS-CoV-2 in the COG-UK database by variant to 18 December 2021. All data are plotted by week of sample collection. Sources: data from GOV.UK Coronavirus (COVID-19) in the UK [17] and COVID-19 Genomics UK (COG-UK) Consortium [18].

### Data

We draw on the integrated national-level SARS-CoV-2 genomic surveillance records of the COVID-19 Genomics UK (COG-UK) Consortium [13]. Lineage counts by local authority area and week of sample collection for England were accessed from the COG-UK website [18] for a 68-week period, September 2020 (epidemiological week 36, ending 5 September) to December 2021 (epidemiological week 50, ending 18 December) (Figure 1B). The data set included geo-coded information on 979,075 SARS-CoV-2 samples assigned to the 309 Lower Tier Local Authority (LTLA) divisions of England. Here, we define the 309 LTLAs according to their most recent (May 2021) status. Information on the lineage of 20,655 samples (2.1%) was either suppressed (1,105) or not recorded (19,550). Of the remaining 958,420 samples, the majority (93.8%) were classified as belonging to the B.1.1.7 and Q (Alpha, 153,405 samples), B.1.617.2 and AY (Delta, 722,133 samples) and B.1.1.529 and BA (Omicron, 23,137 samples) lineages (Table 1). Samples belonging to these lineages form the basis of all our analysis.

**Table 1.** Estimated rate of spatial growth 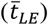 of sample SARS-CoV-2 lineages in England, September 2020–December 2021.

| Variant / lineage | LTLAs (*n*) | Number of detections<sup>1</sup> | Earliest detection<sup>2</sup><br>(*t* = 1) | $\bar{t}_{LE}$ (95% CI)<br>(weeks) |
| --- | --- | --- | --- | --- |
| Alpha (B.1.1.7 and Q) | 307 | 153,405 | 39/2020 (26 Sept.) | 9.90 (9.56, 10.23) |
| Delta (B.1.617.2 and AY) | 307 | 722,133 | 12/2021 (27 March) | 7.40 (7.12, 7.68) |
| B.1.617.2 | 307 | 17,504 | 13/2021 (3 April) | 9.11 (8.48, 9.74) |
| AY.3 | 168 | 620 | 27/2021 (10 July) | 14.12 (13.26, 14.98) |
| AY.4 | 307 | 547,403 | 12/2021 (27 March) | 9.45 (9.24, 9.66) |
| AY.4.1 | 133 | 416 | 25/2021 (26 June) | 9.50 (8.61, 10.40) |
| AY.4.2 | 307 | 77,391 | 25/2021 (26 June) | 6.21 (5.86, 6.57) |
| AY.4.2.1 | 307 | 11,541 | 29/2021 (24 July) | 9.43 (8.91, 9.94) |
| AY.4.3 | 188 | 781 | 19/2021 (15 May) | 19.93 (18.92, 20.93) |
| AY.4.5 | 158 | 742 | 24/2021 (19 June) | 15.06 (13.76, 16.37) |
| AY.5 | 307 | 26,111 | 15/2021 (17 April) | 9.83 (9.38, 10.28) |
| AY.6 | 306 | 17,405 | 17/2021 (1 May) | 8.88 (8.48, 9.27) |
| AY.7 | 276 | 3,627 | 18/2021 (8 May) | 9.02 (8.39, 9.65) |
| AY.8 | 141 | 1,241 | 16/2021 (24 April) | 8.60 (7.90, 9.29) |
| AY.9 | 304 | 10,136 | 14/2021 (10 April) | 10.88 (10.33, 11.42) |
| AY.9.2 | 251 | 1,990 | 28/2021 (17 July) | 10.44 (9.70, 11.17) |
| AY.10 | 118 | 683 | 15/2021 (17 April) | 8.50 (7.96, 9.04) |
| AY.25 | 130 | 486 | 31/2021 (7 Aug.) | 13.02 (12.26, 13.78) |
| AY.33 | 105 | 319 | 25/2021 (26 June) | 17.84 (16.63, 19.05) |
| AY.34 | 268 | 3,088 | 30/2021 (31 July) | 10.88 (10.19, 11.57) |
| AY.36 | 256 | 1,999 | 28/2021 (17 July) | 12.69 (12.05, 13.32) |
| AY.42 | 115 | 338 | 27/2021 (10 July) | 10.08 (8.81, 11.34) |
| AY.43 | 307 | 23,068 | 27/2021 (10 July) | 5.56 (5.27, 5.86) |
| AY.46 | 305 | 9,461 | 21/2021 (29 May) | 11.23 (10.72, 11.74) |
| AY.46.5 | 304 | 8,649 | 23/2021 (12 June) | 10.13 (9.66, 10.61) |
| AY.87 | 172 | 688 | 20/2021 (22 May) | 10.28 (9.28, 11.29) |
| AY.89 | 173 | 544 | 21/2021 (29 May) | 13.97 (13.17, 14.77) |
| AY.90 | 218 | 1,202 | 21/2021 (29 May) | 13.52 (12.54, 14.50) |
| AY.98 | 307 | 22,465 | 22/2021 (5 June) | 6.34 (5.99, 6.70) |
| AY.98.1 | 172 | 621 | 25/2021 (26 June) | 15.77 (14.80, 16.75) |
| AY.109 | 116 | 441 | 25/2021 (26 June) | 17.59 (16.67, 18.52) |
| AY.111 | 296 | 4,649 | 21/2021 (29 May) | 15.25 (14.46, 16.04) |
| AY.120 | 306 | 18,400 | 20/2021 (22 May) | 6.56 (6.16, 6.97) |
| AY.122 | 302 | 5,018 | 21/2021 (29 May) | 13.80 (13.07, 14.53) |
| AY.124 | 119 | 424 | 25/2021 (26 June) | 12.99 (11.93, 14.05) |
| AY.125 | 162 | 534 | 26/2021 (3 July) | 16.65 (15.79, 17.51) |
| Omicron (B.1.1.529 and BA) | 304 | 23,137 | 47/2021 (27 Nov.)<sup>3</sup> | 2.63 (2.56, 2.71)<sup>4</sup> |
| Average | 233 | 46,450 |  | 11.27 (10.03, 12.50) |

<sup>1</sup> Excludes 1,105 detections for which lineage data is suppressed and 19,550 detections for which lineage data is not available. <sup>2</sup> Epidemiological week/year, with the last day of the week given in parentheses. <sup>3</sup> Excludes a lone detection in week 43 (30 October). <sup>4</sup> Indexed to week 47; $\bar{t}_{LE}$ = 6.62 (6.53, 6.70) when indexed to week 43.

## Methods

To assess the spatial growth rate of a given SARS-CoV-2 lineage, we draw on the *swash– backwash model of the single epidemic wave* [12]. In essence, the model represents a spatial derivative of the generic *SIR* mass action models of infectious disease transmission [19]. Using the binary (presence/absence) of a disease, the model (i) allows the disaggregation of an infection wave into phases of spatial expansion and retreat and (ii) provides a means of measuring the phase transitions of geographical units from susceptible *S*, through infective *I* to recovered *R* status. See, for example, Smallman-Raynor and Cliff [20] and Smallman-Raynor, *et al*. [21].

### Measuring the spatial growth rate

Full details of the modelling procedure are outlined by Cliff and Haggett [12]. For the purposes of the present analysis, we focus on the spatial expansion phase (that is, the change of state from *S* to *I* across a set geographical units) for a given SARS-CoV-2 lineage. Specifically, let the first week in which the lineage was detected in England be coded as *t* = 1. Subsequent weeks were then coded serially as *t* = 2, 3, …, *T*, where *T* is the number of weekly periods from the beginning to the end of the detected occurrence of the lineage. For any given geographical unit, we refer to the first week in which the lineage was detected as the *leading edge* (*LE*) of the infection wave. The average time (in weeks) to the detection of the lineage across the set of units can then be defined by a time-weighted mean, 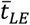, of the form

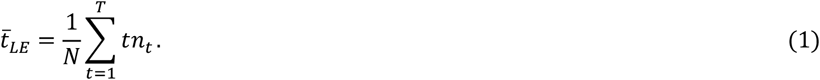

Here, *n*_*t*_ is the number of units whose leading edge, *LE*, occurred in week *t* and *N* = ∑ *n*_*t*_. Formed in this manner, SARS-CoV-2 lineages with relatively *high* rates of spatial expansion (or rapidly developing *LE*) take on relatively *low* values of 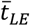 (i.e. short average times to detection). Conversely, lineages with relatively *low* rates of spatial expansion (or slowly developing *LE*) take on relatively *high* values of 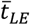 (i.e. long average times to detection).

### Application of the model

Equation (1) was used to estimate the spatial growth rate of sample SARS-CoV-2 lineages for which the earliest detection in England occurred in the time period covered by the dataset (September 2020–December 2021) and for which substantial geographical spread had been documented. To ensure the inclusion of sufficient observations for geographical analysis, the sample was limited to lineages that had been detected in at least one-third of the 309 LTLAs by December 2021. Based on these criteria, the sample consisted of 37 lineages. Summary details of the sample, including the number of LTLAs in which each lineage was detected, the total count of detections over the study period, and the earliest date of detection, are provided in Table 1.

For each lineage, equation (1) was fitted with *t* = 1 set to the week of earliest detection in Table 1. In the instance of Omicron, retrospective analysis has identified a lone detection of the BA.1 lineage in epidemiological week 43 of 2021 (week ending 30 October), four weeks prior to the subsequent detection and apparent onset of widespread transmission of the variant in epidemiological week 47 (week ending 27 November). For the purposes of the present analysis, we set week 47 as *t* = 1 for Omicron, but we also report the computed value of 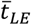 based on the earlier detection in week 43. Finally, we exclude two LTLAs (City of London and Isles of Scilly) from all analysis on account of the suppression of lineage data due to their small populations. Data analysis was performed in Minitab^®^17 (Minitab Inc., Pennsylvania) and data mapping in QGIS 3.10.14-A Coruña (QGIS.org) using Local Authority Districts (May 2021) UK and Regions (December 2020) EN shapefiles from the Office for National Statistics (ONS) [22].

## Results

Table 1 confirms that the 37 sample lineages were geographically extensive in their transmission, with 29 having been detected in >150 LTLAs, 21 in >250 LTLAs, 16 in >300 LTLAs, and nine in the complete set of 307 LTLAs under examination. The majority (23) were associated with >1,000 detections, 13 with >10,000 detections and three with >100,000 detections. Delta (B.1.617.2 and AY) was the most common lineage (722,133 detections) and AY.4 the most common sublineage (547,403 detections), with AY lineages accounting for 33 of the spread events under examination. In turn, the majority of lineages emerged (as judged by the date of earliest detections) in the spring and summer of 2021, as the Delta infection wave was evolving both domestically and internationally.

### Spatial growth curves and leading edge (*LE*) maps

The upper graphs in Figure 2 plot the count of LTLAs by week of earliest detection of the Alpha (B.1.1.7 and Q), Delta (B.1.617.2 and AY) and Omicron (B.1.1.529 and BA) variants, where weeks are indexed to the earliest detection of the respective variants (Table 1). The lower graphs are spatial growth curves, formed by replotting the information in the upper graphs as a cumulative proportion of LTLAs. Average curves for the set of sample lineages in Table 1 are shown for reference.

**Figure 2.**
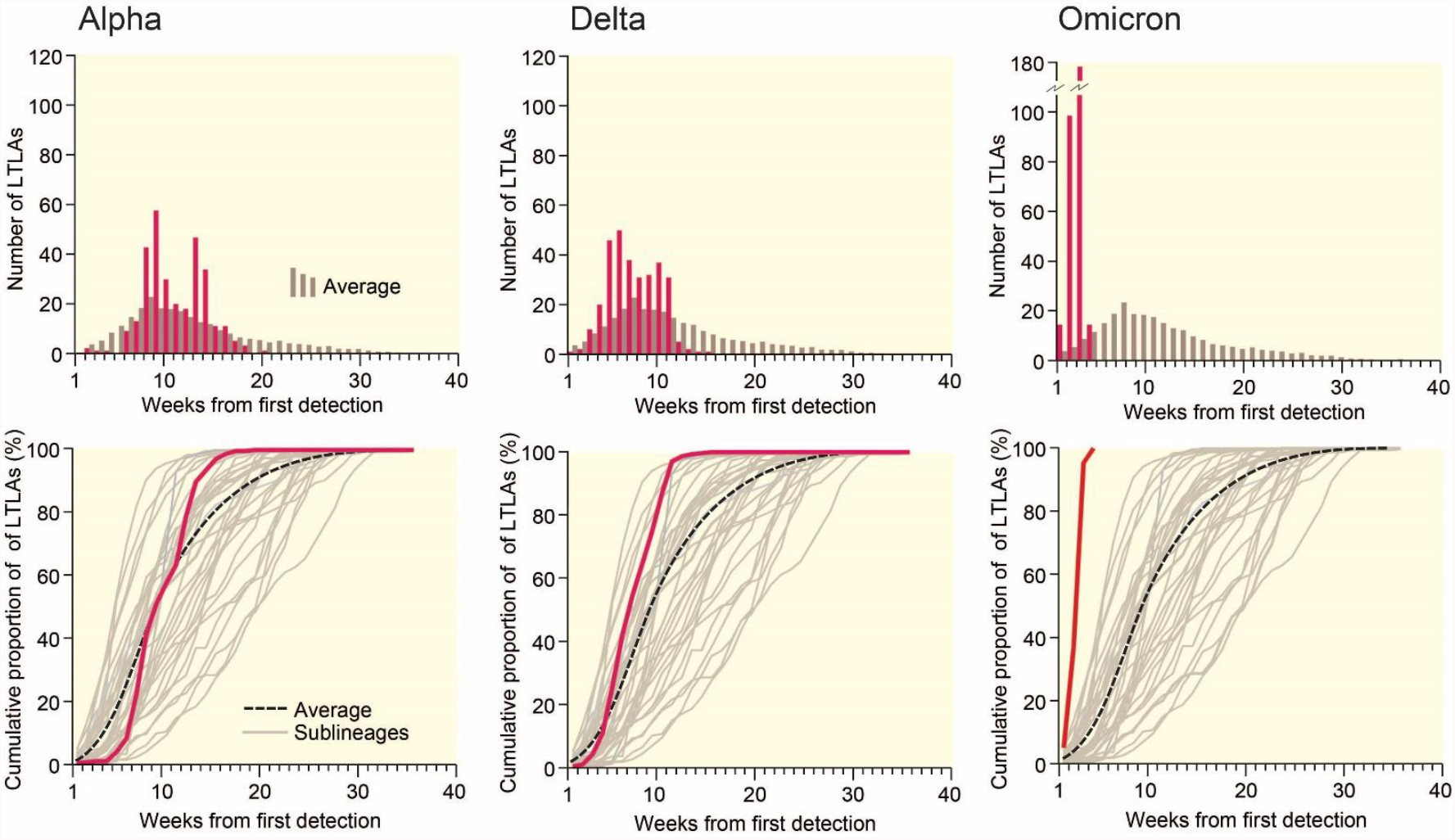
Spatial leading edges (*LE*) of the Alpha (B.1.1.7 and Q), Delta (B.1.617.2 and AY) and Omicron (B.1.1529 and BA) variants in England, September 2020–December 2021. The graphs plot, on a weekly basis, the non-cumulative count (upper) and cumulative proportion (lower) of LTLAs in which each of the three variants was first detected. The horizontal (time) axes are indexed to the epidemiological week of first detection (*t* = 1) of the corresponding variant. Average curves, formed across the set of sample lineages in Table 1, are plotted for reference.

Together, the graphs in Figure 2 portray the temporal development of the spatial leading edges (*LE*) for each variant. The geographical expression of these *LE* is captured by the choropleth maps in Figure 3 which plot the week of earliest detection of each variant in the set of LTLAs. The sequentially more rapid spatial growth of the variants (Alpha → Delta → Omicron) is evidenced by the sequentially steeper spatial growth curves (Figure 2) and the sequentially shorter periods to earliest detection (Figure 3). The latter feature is emphasized when earliest detections are formed as regional averages in Figure 4.

**Figure 3.**
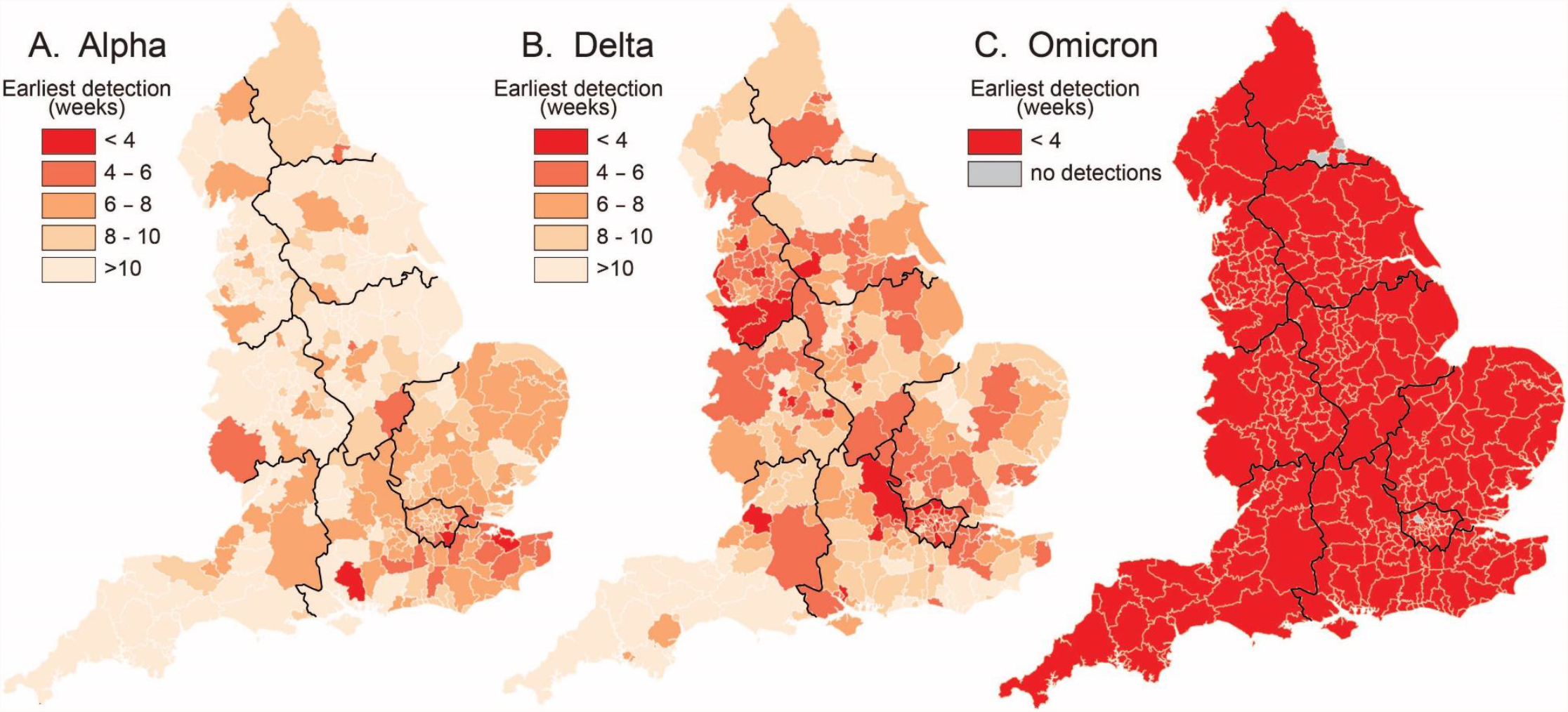
Spatial leading edges (*LE*) of the Alpha (B.1.1.7 and Q), Delta (B.1.617.2 and AY) and Omicron (B.1.1529 and BA) variants in the LTLAs of England, September 2020–December 2021. Maps are indexed to the epidemiological week of first detection (= week 1) of the corresponding variant and plot the number of weeks to first detection in each LTLA.

**Figure 4.**
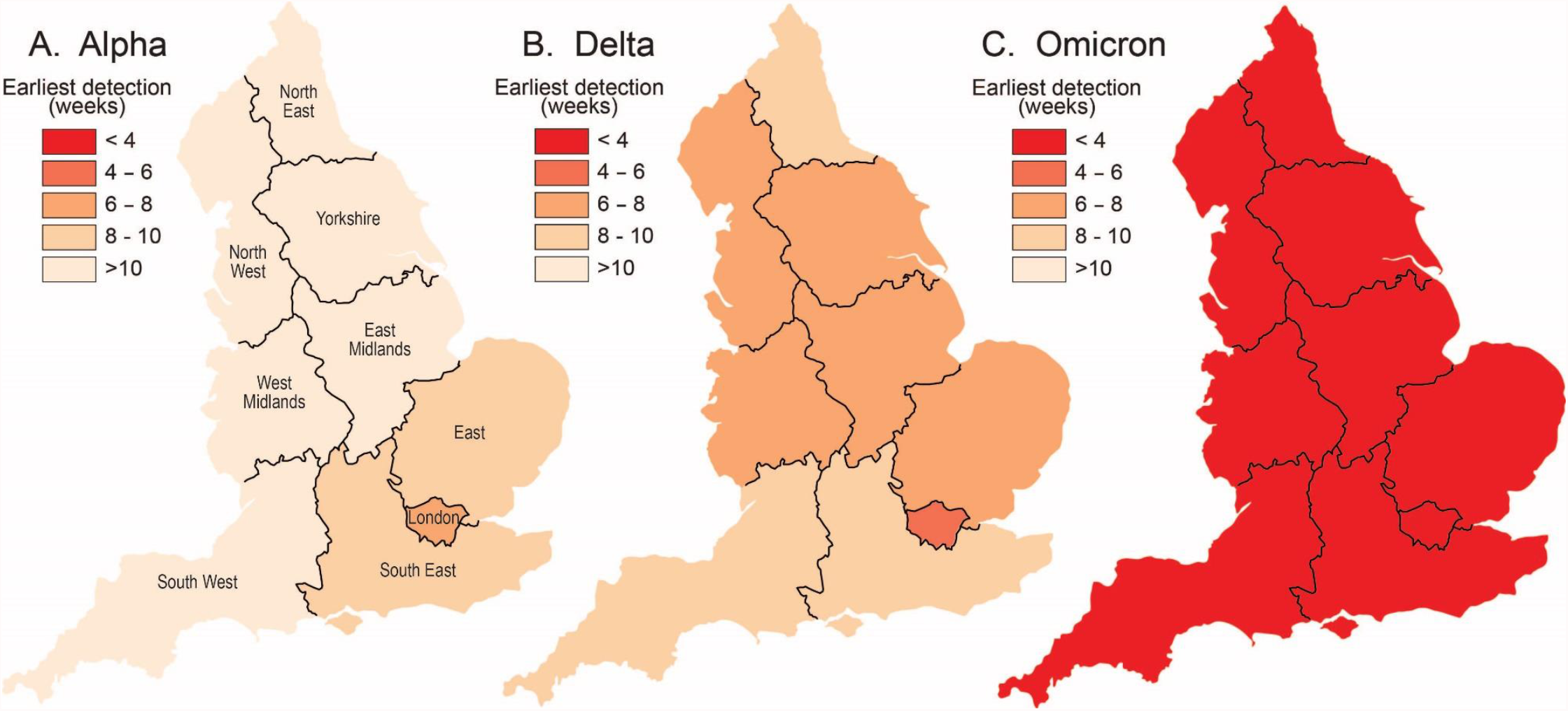
Spatial leadings edges (*LE*) of the Alpha (B.1.1.7 and Q), Delta (B.1.617.2 and AY) and Omicron (B.1.1529 and BA) variants in the nine standard regions of England, September 2020–December 2021. Maps plot the average time (in weeks) to first detection of a given variant in each regional subset of LTLAs.

### Rates of spatial growth 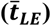

The right-hand column in Table 1 summarizes the results of the application of equation (1) to each of the sample lineages. Computed values of 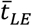 and associated 95% confidence intervals (95% CI) are given, along with an overall average value of 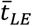 for the entire sample. As noted above, lineages with relatively *high* rates of spatial expansion (or rapidly developing *LE*) are represented by relatively *low* values of 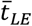 (i.e. short average times to detection), while lineages with relatively *low* rates of spatial expansion (or slowly developing *LE*) take on relatively *high* values of 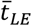 (i.e. long average times to detection). In this manner, the table confirms the sequential increase in the spatial growth rate for Alpha, Delta and Omicron. On average, the earliest detection of the Alpha variant in a given LTLA occurred at 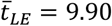 (95% CI: 9.56, 10.23) weeks after the earliest sampled detection in England. This reduced to 7.40 (95% CI: 7.12, 7.68) weeks for Delta and 2.63 (95% CI: 2.56, 2.71) weeks for Omicron.

### Delta AY lineages

Figure 5 is based on the information in Table 1 and plots the values of 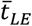 for B.1.617.2 and AY lineages in order, from the lowest (left, high values of 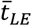) to the highest (right, low values of 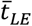) rates of spatial growth. Values are plotted on an inverted vertical scale to facilitate interpretation. The average value of 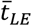, formed across the sample set of lineages in Table 1, is indicated for reference as are the 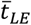 for the Alpha (B.1.1.7 and Q), Delta (B.1.617.2 and AY) and Omicron (B.1.1.529 and BA) variants. Spatial growth curves, formed in the manner of Figure 2, are plotted for a sample of 20 AY lineages with relatively high and low rates of spatial growth in Figure 6.

**Figure 5.**
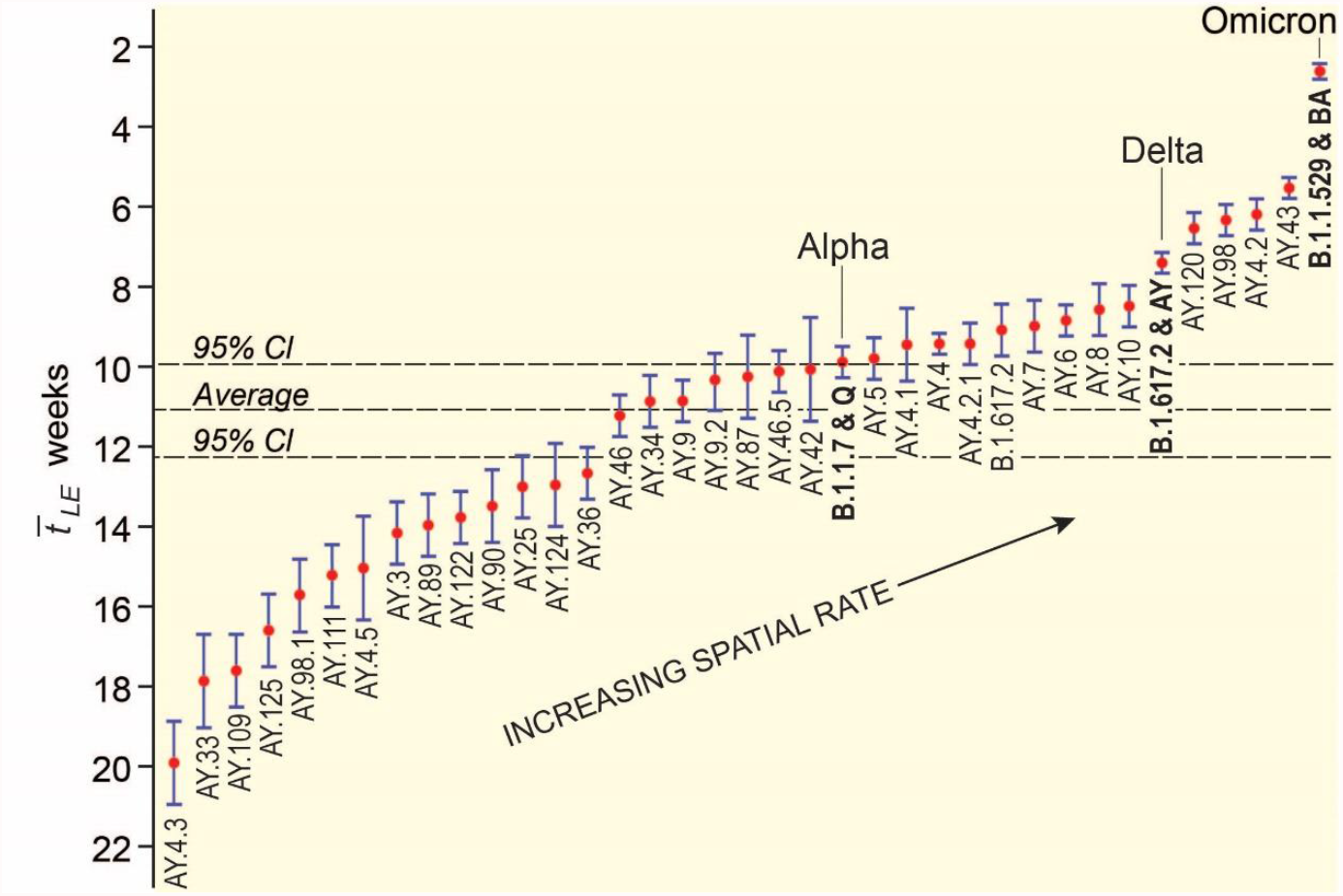
Estimated rate of spatial growth of sample SARS-CoV-2 lineages in England, September 2020–December 2021. The graph plots values of 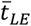 and associated 95% CI from Table 1. Values are ordered from the lowest (left, high values of 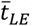) to the highest (right, low values of 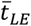) rates of spatial growth. Values are plotted on an inverted vertical scale to facilitate interpretation. The average value of 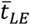 for the sample is shown for reference.

**Figure 6.**
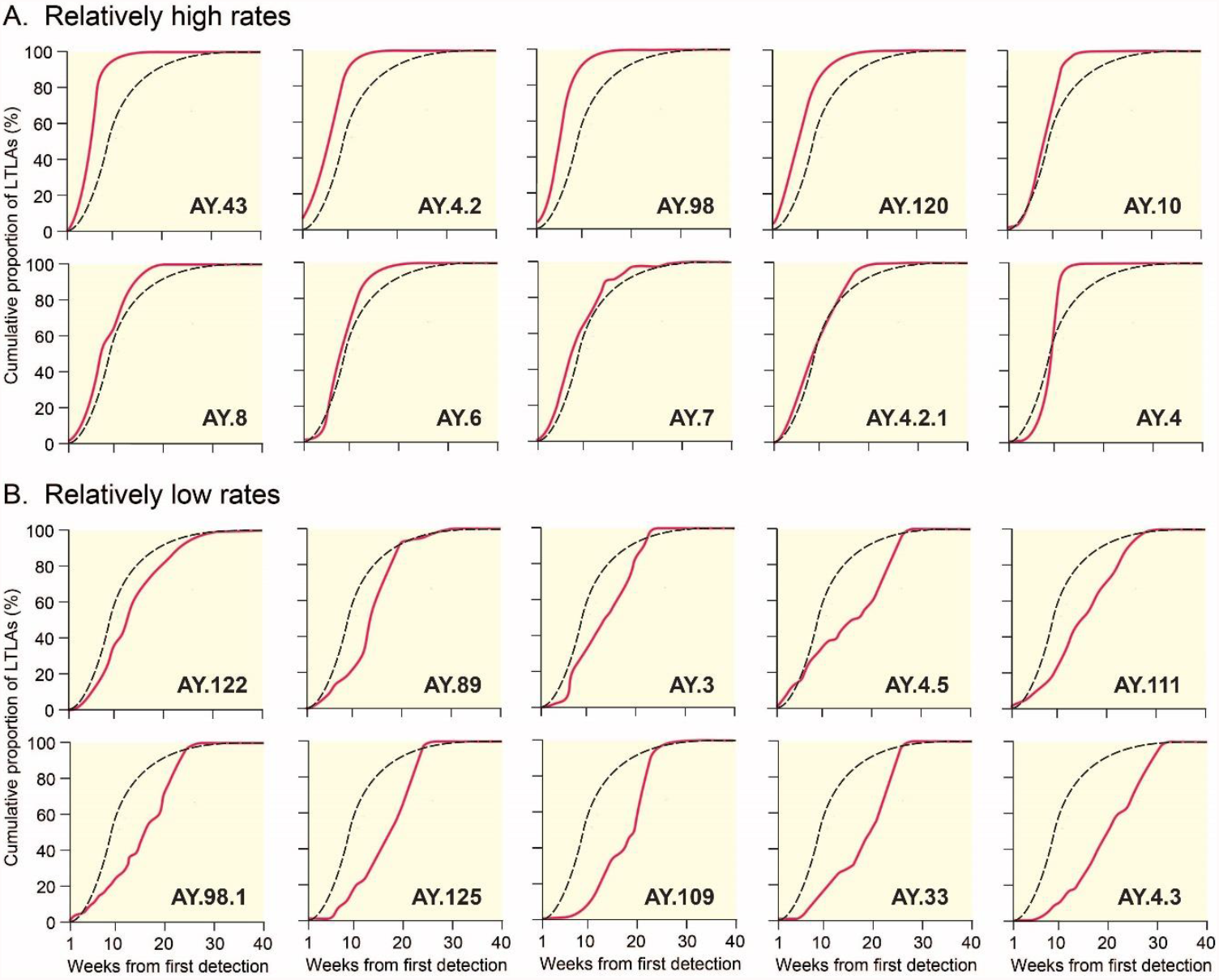
Spatial growth curves for sample Delta sublineages in England, March–December 2021. Curves have been formed in the manner of the lower graphs in Figure 2, with the average curve plotted for reference. Lineages are ordered according to the values of 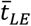 in Table 1 and are defined as having relatively high (i.e. low values of 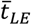; upper graphs, A) and relatively low (i.e. high values of 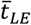; lower graphs, B) rates of spatial growth.

There is a 7.6-fold difference in the range of values of 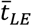 in Figure 5, from Delta AY.4.3 with the lowest spatial growth rate (19.93 weeks) to Omicron with the highest (2.63 weeks). A group of four AY lineages (AY.4.2, AY.43, AY.98 and AY.120), first detected in the period from mid-May to mid-July 2021, are positioned between Delta and Omicron in Figure 5 and display rates of spatial growth that are significantly higher (as judged by 95% CI) than the aggregate rate for the Delta variant. In contrast, the overwhelming majority of AY lineages display statistically lower – in many instances substantially lower – spatial growth rates (as judged by 95% CI) than the aggregate rate for the Delta variant.

## Discussion

Recent experience has underscored the importance of the ongoing tracking, monitoring and analysis of emerging SARS-CoV-2 lineages with a view to mitigating the impacts of the COVID-19 pandemic [23]. We have used a robust model of spatial epidemiological analysis to estimate the rate of spatial growth of multiple lineages of the virus in England over a 68-week period, September 2020–December 2021. We have shown that the Alpha, Delta and Omicron variants took an average of 9.90 weeks, 7.40 weeks and 2.63 weeks, respectively, to reach the set of LTLAs under examination (Table 1 and Figure 5). Expressed in relative terms, the leading spatial edges were 1.34× faster (Delta vs. Alpha), 2.81× faster (Omicron vs. Delta), and 3.76× faster (Omicron vs. Alpha). Our estimates scale to the approximate length of time that Alpha (12 weeks), Delta (8 weeks) and Omicron (3 weeks) took to establish themselves as the dominant variants in England [18], and are consistent with evidence for the fitness advantage of Delta over Alpha and Omicron over Delta [11, 24, 25].

Of the 121 Delta AY lineages detected in England to December 2021 and included in the genomic surveillance records of the COG-UK Consortium, 33 lineages met the geographical criterion for inclusion in the current analysis. In interpreting the results for these lineages, we note that AY designations are phylogenetically defined and do not necessarily denote any fundamental biological differences between the lineages [26]. Moreover, results of the type documented in this paper are context dependent and cannot be interpreted as evidence of a change in biological transmissibility, immune escape or other fitness advantage. Subject to these caveats, we have identified four AY lineages (AY.4.2, AY.43, AY.98 and AY.120) for which the rate of spatial growth exceeded the aggregate rate for the Delta variant. These lineages had been detected in all (AY.4.2, AY.43 and AY.98) or most (AY.120) of the local authority areas under investigation, and each had been associated with considerably more than 10,000 detections (Table 1). Table 2 summarizes the global status of these four lineages as of 9 January 2022. With the exception of the AY.43 lineage, which was prevalent in a number of European countries and associated with >267,000 detections worldwide, the majority of detections of these lineages originated from the United Kingdom.

**Table 2.** Worldwide detection of sample Delta AY lineages with relatively high estimated rates of spatial growth (status: 9 January 2022).

| Variant | Countries | Global prevalence (%) | Number of detections | Predominant countries (proportion of global detections) |
| --- | --- | --- | --- | --- |
| AY.4.2 | 52 | 1 | 82,038 | UK (90%), Denmark (4%), Germany (1%), Poland (1%), France (1%) |
| AY.43 | 128 | 4 | 267,426 | Germany (15%), UK (13%), Denmark (13%), France (12%), Belgium (5%) |
| AY.98 | 66 | 1 | 41,507 | UK (91%), USA (2%), Germany (1%), Denmark (1%), Ireland (1%) |
| AY.120 | 74 | 1 | 30,320 | UK (85%), India (3%), USA (3%), Germany (2%), France (1%) |

*Sources:* data from cov-lineages.org [27] and Latif, *et al.* [28–31].

Our findings for the AY.4.2 and AY.43 lineages are consistent with their respective designations by the UK Health Security Agency as a distinct variant under investigation (VUI-21OCT-01) and a variant of concern [32, 33]. Preliminary investigations have indicated the AY.4.2 lineage to be associated with a higher growth rate and a higher household secondary attack rate, but with no significant reduction in vaccine effectiveness, as compared to the parent lineage [32, 34]. It remains to be established, however, whether the higher growth rate is due to enhanced transmissibility or is context-dependent [32, 35, 36]. Similarly, the status of the AY.43 lineage in terms of transmission advantage and/or immune escape remains to be determined, although further investigation is merited as new AY.43 sublineages have recently been reported from Brazil [37]. Finally, our identification of a rapid rate of spatial growth for the AY.98 and AY.120 lineages, approximating the estimated rates for AY.4.2 and AY.43, is noteworthy. Whilst very little has been documented on the epidemiological facets of these lineages, both have been identified in a number of countries in Europe and elsewhere (Table 2) and would seem to merit further investigation on the basis of the findings presented here.

With the foregoing exceptions, our analysis has shown that many emerging AY lineages in England in the spring and summer of 2021 were associated with spatial growth rates that were lower (in some instances, substantially lower) than the aggregate rate for the Delta variant (Table 1 and Figure 5). Multiple biological (e.g. reduced infectivity or transmissibility) and contextual (e.g. progressive expansion of the national COVID-19 vaccination programme) factors may account for this observation. Importantly, there is no evidence of a temporal trend in the observed rates of spatial growth that would be suggestive of either (i) a biological selection pressure in favour of a growth advantage of emerging lineages or (ii) a progressive contextual effect in the form of, for example, increasing levels of vaccination coverage or natural immunity that would serve to retard growth rates.

Our estimates of 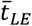 in Table 1 and Figure 5 are contingent on a range of public health interventions that operated to limit the spread of SARS-CoV-2. These interventions included: a tier system of local lockdown in October 2020; two periods of national lockdown (November–December 2020 and January–March 2021) and a phased lifting of national restrictions in the period to July 2021; a phased rollout of the national COVID-19 vaccination programme from December 2020; and the implementation of ‘Plan B’ control guidelines against the Omicron variant in December 2021 [38, 39]. Our results are also subject to the limitations of the available lineage data. Although the COG-UK Consortium genomic surveillance data are recognized for their extent and reliability [40], the data are formed as a sample of positive SARS-CoV-2 test results and are subject to the limitations and biases of sample data. In this context, we note that the cumulative coverage of the COG-UK records for England was estimated at 13.7% of people with positive SARS-CoV-2 test results to October 2021 [41]. Finally, our results are subject to the underpinning assumptions of the analytical procedure. In particular, the computation of 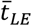 is dependent on the specification of the index week (i.e. the week that a given lineage was first detected in England) and the degree to which this reflects the date of actual emergence of the lineage in England. The extent to which the sample data accurately track the spatial expansion of the *LE* for a given lineage, the variable contributions of international travel- and community-related transmission to the development of the *LE*, and the geographical starting point(s) of a given lineage in the national transmission network, will also have influenced our results in unknown ways. For example, the early involvement and high degree of geographical connectivity of London and the South East may have served to accelerate the spatial transmission of the Alpha variant in the latter months of 2020 [14]. The observed rapid spread of the Delta variant may reflect international importations and onwards transmission from multiple different geographical locations in the spring of 2021 [15, 42], whilst early cases of the Omicron variant were observed in highly connected regions at a time of reduced non-pharmaceutical interventions in November and December 2021 [43].

Further insights into the spatial growth and decay of SARS-CoV-2 lineages may be gained by application of the full swash-backwash model, but this is dependent on the substantial spatial retreat of any given lineage from the population. Here we note that, with the exception of AY.10 (last detected in July 2021) and AY.8 and Alpha (B.1.1.7 and Q) (both last detected in August/September 2021), there is evidence of the circulation of all the lineages included in Table 1 in the weeks to December 2021.

We have demonstrated, for the first time, a robust method for assessing and comparing the rate of spatial growth of multiple SARS-CoV-2 lineages in a set of geographical areas. We suggest that this approach represents a potentially valuable adjunct to routine assessments of the growth of emerging SARS-CoV-2 lineages in a defined population.

## Data Availability

The data that support the findings of this study are available at Wellcome Sanger Institute COVID-19 Genomic Surveillance (https://covid19.sanger.ac.uk/lineages/raw).

https://geoportal.statistics.gov.uk/

### Appendix: The COVID-19 Genomics UK (COG-UK) Consortium

**Funding acquisition, Leadership and supervision, Metadata curation, Project administration, Samples and logistics, Sequencing and analysis, Software and analysis tools, and Visualisation:**

Samuel C Robson ^13, 84^

**Funding acquisition, Leadership and supervision, Metadata curation, Project administration, Samples and logistics, Sequencing and analysis, and Software and analysis tools:**

Thomas R Connor ^11, 74^ and Nicholas J Loman ^43^

**Leadership and supervision, Metadata curation, Project administration, Samples and logistics, Sequencing and analysis, Software and analysis tools, and Visualisation:**

Tanya Golubchik ^5^

**Funding acquisition, Leadership and supervision, Metadata curation, Samples and logistics, Sequencing and analysis, and Visualisation:**

Rocio T Martinez Nunez ^46^

**Funding acquisition, Leadership and supervision, Project administration, Samples and logistics, Sequencing and analysis, and Software and analysis tools:**

David Bonsall ^5^

**Funding acquisition, Leadership and supervision, Project administration, Sequencing and analysis, Software and analysis tools, and Visualisation:**

Andrew Rambaut ^104^

**Funding acquisition, Metadata curation, Project administration, Samples and logistics, Sequencing and analysis, and Software and analysis tools:**

Luke B Snell ^12^

**Leadership and supervision, Metadata curation, Project administration, Samples and logistics, Software and analysis tools, and Visualisation:**

Rich Livett ^116^

**Funding acquisition, Leadership and supervision, Metadata curation, Project administration, and Samples and logistics:**

Catherine Ludden ^20, 70^

**Funding acquisition, Leadership and supervision, Metadata curation, Samples and logistics, and Sequencing and analysis:**

Sally Corden ^74^ and Eleni Nastouli ^96, 95, 30^

**Funding acquisition, Leadership and supervision, Metadata curation, Sequencing and analysis, and Software and analysis tools:**

Gaia Nebbia ^12^

**Funding acquisition, Leadership and supervision, Project administration, Samples and logistics, and Sequencing and analysis:**

Ian Johnston ^116^

**Leadership and supervision, Metadata curation, Project administration, Samples and logistics, and Sequencing and analysis:**

Katrina Lythgoe ^5^, M. Estee Torok ^19, 20^ and Ian G Goodfellow ^24^

**Leadership and supervision, Metadata curation, Project administration, Samples and logistics, and Visualisation:**

Jacqui A Prieto ^97, 82^ and Kordo Saeed ^97, 83^

**Leadership and supervision, Metadata curation, Project administration, Sequencing and analysis, and Software and analysis tools:**

David K Jackson ^116^

**Leadership and supervision, Metadata curation, Samples and logistics, Sequencing and analysis, and Visualisation:**

Catherine Houlihan ^96, 94^

**Leadership and supervision, Metadata curation, Sequencing and analysis, Software and analysis tools, and Visualisation:**

Dan Frampton ^94, 95^

**Metadata curation, Project administration, Samples and logistics, Sequencing and analysis, and Software and analysis tools:**

William L Hamilton ^19^ and Adam A Witney ^41^

**Funding acquisition, Samples and logistics, Sequencing and analysis, and Visualisation:**

Giselda Bucca ^101^

**Funding acquisition, Leadership and supervision, Metadata curation, and Project administration:**

Cassie F Pope ^40, 41^

**Funding acquisition, Leadership and supervision, Metadata curation, and Samples and logistics:**

Catherine Moore ^74^

**Funding acquisition, Leadership and supervision, Metadata curation, and Sequencing and analysis:**

Emma C Thomson ^53^

**Funding acquisition, Leadership and supervision, Project administration, and Samples and logistics:**

Ewan M Harrison ^116, 102^

**Funding acquisition, Leadership and supervision, Sequencing and analysis, and Visualisation:**

Colin P Smith ^101^

**Leadership and supervision, Metadata curation, Project administration, and Sequencing and analysis:**

Fiona Rogan ^77^

**Leadership and supervision, Metadata curation, Project administration, and Samples and logistics:**

Shaun M Beckwith ^6^, Abigail Murray ^6^, Dawn Singleton ^6^, Kirstine Eastick ^37^, Liz A Sheridan ^98^, Paul Randell ^99^, Leigh M Jackson ^105^, Cristina V Ariani ^116^ and Sónia Gonçalves ^116^

**Leadership and supervision, Metadata curation, Samples and logistics, and Sequencing and analysis:**

Derek J Fairley ^3, 77^, Matthew W Loose ^18^ and Joanne Watkins ^74^

**Leadership and supervision, Metadata curation, Samples and logistics, and Visualisation:**

Samuel Moses ^25, 106^

**Leadership and supervision, Metadata curation, Sequencing and analysis, and Software and analysis tools:**

Sam Nicholls ^43^, Matthew Bull ^74^ and Roberto Amato ^116^

**Leadership and supervision, Project administration, Samples and logistics, and Sequencing and analysis:**

Darren L Smith ^36, 65, 66^

**Leadership and supervision, Sequencing and analysis, Software and analysis tools, and Visualisation:**

David M Aanensen ^14, 116^ and Jeffrey C Barrett ^116^

**Metadata curation, Project administration, Samples and logistics, and Sequencing and analysis:**

Dinesh Aggarwal ^20, 116, 70^, James G Shepherd ^53^, Martin D Curran ^71^ and Surendra Parmar ^71^

**Metadata curation, Project administration, Sequencing and analysis, and Software and analysis tools:**

Matthew D Parker ^109^

**Metadata curation, Samples and logistics, Sequencing and analysis, and Software and analysis tools:**

Catryn Williams ^74^

**Metadata curation, Samples and logistics, Sequencing and analysis, and Visualisation:**

Sharon Glaysher ^68^

**Metadata curation, Sequencing and analysis, Software and analysis tools, and Visualisation:**

Anthony P Underwood ^14, 116^, Matthew Bashton ^36, 65^, Nicole Pacchiarini ^74^, Katie F Loveson ^84^ and

Matthew Byott ^95, 96^

**Project administration, Sequencing and analysis, Software and analysis tools, and Visualisation:**

Alessandro M Carabelli ^20^

**Funding acquisition, Leadership and supervision, and Metadata curation:**

Kate E Templeton ^56, 104^

**Funding acquisition, Leadership and supervision, and Project administration:**

Thushan I de Silva ^109^, Dennis Wang ^109^, Cordelia F Langford ^116^ and John Sillitoe ^116^

**Funding acquisition, Leadership and supervision, and Samples and logistics:**

Rory N Gunson ^55^

**Funding acquisition, Leadership and supervision, and Sequencing and analysis:**

Simon Cottrell ^74^, Justin O’Grady ^75, 103^ and Dominic Kwiatkowski ^116, 108^

**Leadership and supervision, Metadata curation, and Project administration:**

Patrick J Lillie ^37^

**Leadership and supervision, Metadata curation, and Samples and logistics:**

Nicholas Cortes ^33^, Nathan Moore ^33^, Claire Thomas ^33^, Phillipa J Burns ^37^, Tabitha W Mahungu ^80^ and Steven Liggett ^86^

**Leadership and supervision, Metadata curation, and Sequencing and analysis:**

Angela H Beckett ^13, 81^ and Matthew TG Holden ^73^

**Leadership and supervision, Project administration, and Samples and logistics:**

Lisa J Levett ^34^, Husam Osman ^70, 35^ and Mohammed O Hassan-Ibrahim ^99^

**Leadership and supervision, Project administration, and Sequencing and analysis:**

David A Simpson ^77^

**Leadership and supervision, Samples and logistics, and Sequencing and analysis:**

Meera Chand ^72^, Ravi K Gupta ^102^, Alistair C Darby ^107^ and Steve Paterson ^107^

**Leadership and supervision, Sequencing and analysis, and Software and analysis tools:**

Oliver G Pybus ^23^, Erik M Volz ^39^, Daniela de Angelis ^52^, David L Robertson ^53^, Andrew J Page ^75^ and Inigo Martincorena ^116^

**Leadership and supervision, Sequencing and analysis, and Visualisation:**

Louise Aigrain ^116^ and Andrew R Bassett ^116^

**Metadata curation, Project administration, and Samples and logistics:**

Nick Wong ^50^, Yusri Taha ^89^, Michelle J Erkiert ^99^ and Michael H Spencer Chapman ^116, 102^

**Metadata curation, Project administration, and Sequencing and analysis:**

Rebecca Dewar ^56^ and Martin P McHugh ^56, 111^

**Metadata curation, Project administration, and Software and analysis tools:**

Siddharth Mookerjee ^38, 57^

**Metadata curation, Project administration, and Visualisation:**

Stephen Aplin ^97^, Matthew Harvey ^97^, Thea Sass ^97^, Helen Umpleby ^97^ and Helen Wheeler ^97^

**Metadata curation, Samples and logistics, and Sequencing and analysis:**

James P McKenna ^3^, Ben Warne ^9^, Joshua F Taylor ^22^, Yasmin Chaudhry ^24^, Rhys Izuagbe ^24^, Aminu S Jahun ^24^, Gregory R Young ^36, 65^, Claire McMurray ^43^, Clare M McCann ^65, 66^, Andrew Nelson ^65, 66^ and

Scott Elliott ^68^

**Metadata curation, Samples and logistics, and Visualisation:**

Hannah Lowe ^25^

**Metadata curation, Sequencing and analysis, and Software and analysis tools:**

Anna Price ^11^, Matthew R Crown ^65^, Sara Rey ^74^, Sunando Roy ^96^ and Ben Temperton ^105^

**Metadata curation, Sequencing and analysis, and Visualisation:**

Sharif Shaaban ^73^ and Andrew R Hesketh ^101^

**Project administration, Samples and logistics, and Sequencing and analysis:**

Kenneth G Laing ^41^, Irene M Monahan ^41^ and Judith Heaney ^95, 96, 34^

**Project administration, Samples and logistics, and Visualisation:**

Emanuela Pelosi ^97^, Siona Silviera ^97^ and Eleri Wilson-Davies ^97^

**Samples and logistics, Software and analysis tools, and Visualisation:**

Helen Fryer ^5^

**Sequencing and analysis, Software and analysis tools, and Visualization:**

Helen Adams ^4^, Louis du Plessis ^23^, Rob Johnson ^39^, William T Harvey ^53, 42^, Joseph Hughes ^53^, Richard J Orton ^53^, Lewis G Spurgin ^59^, Yann Bourgeois ^81^, Chris Ruis ^102^, Áine O’Toole ^104^, Marina Gourtovaia ^116^ and Theo Sanderson ^116^

**Funding acquisition, and Leadership and supervision:**

Christophe Fraser ^5^, Jonathan Edgeworth ^12^, Judith Breuer ^96, 29^, Stephen L Michell ^105^ and John A Todd ^115^

**Funding acquisition, and Project administration:**

Michaela John ^10^ and David Buck ^115^

**Leadership and supervision, and Metadata curation:**

Kavitha Gajee ^37^ and Gemma L Kay ^75^

**Leadership and supervision, and Project administration:**

Sharon J Peacock ^20, 70^ and David Heyburn ^74^

**Leadership and supervision, and Samples and logistics:**

Katie Kitchman ^37^, Alan McNally ^43, 93^, David T Pritchard ^50^, Samir Dervisevic ^58^, Peter Muir ^70^, Esther Robinson ^70, 35^, Barry B Vipond ^70^, Newara A Ramadan ^78^, Christopher Jeanes ^90^, Danni Weldon ^116^, Jana Catalan ^118^ and Neil Jones ^118^

**Leadership and supervision, and Sequencing and analysis:**

Ana da Silva Filipe ^53^, Chris Williams ^74^, Marc Fuchs ^77^, Julia Miskelly ^77^, Aaron R Jeffries ^105^, Karen Oliver ^116^ and Naomi R Park ^116^

**Metadata curation, and Samples and logistics:**

Amy Ash ^1^, Cherian Koshy ^1^, Magdalena Barrow ^7^, Sarah L Buchan ^7^, Anna Mantzouratou ^7^, Gemma Clark ^15^, Christopher W Holmes ^16^, Sharon Campbell ^17^, Thomas Davis ^21^, Ngee Keong Tan ^22^, Julianne R Brown ^29^, Kathryn A Harris ^29, 2^, Stephen P Kidd ^33^, Paul R Grant ^34^, Li Xu-McCrae ^35^, Alison Cox ^38, 63^, Pinglawathee Madona ^38, 63^, Marcus Pond ^38, 63^, Paul A Randell ^38, 63^, Karen T Withell ^48^, Cheryl Williams ^51^, Clive Graham ^60^, Rebecca Denton-Smith ^62^, Emma Swindells ^62^, Robyn Turnbull ^62^, Tim J Sloan ^67^, Andrew Bosworth ^70, 35^, Stephanie Hutchings ^70^, Hannah M Pymont ^70^, Anna Casey ^76^, Liz Ratcliffe ^76^, Christopher R Jones ^79, 105^, Bridget A Knight ^79, 105^, Tanzina Haque ^80^, Jennifer Hart ^80^, Dianne Irish-Tavares ^80^, Eric Witele ^80^, Craig Mower ^86^, Louisa K Watson ^86^, Jennifer Collins ^89^, Gary Eltringham ^89^, Dorian Crudgington ^98^, Ben Macklin ^98^, Miren Iturriza-Gomara ^107^, Anita O Lucaci ^107^ and Patrick C McClure ^113^

**Metadata curation, and Sequencing and analysis:**

Matthew Carlile ^18^, Nadine Holmes ^18^, Christopher Moore ^18^, Nathaniel Storey ^29^, Stefan Rooke ^73^, Gonzalo Yebra ^73^, Noel Craine ^74^, Malorie Perry ^74^, Nabil-Fareed Alikhan ^75^, Stephen Bridgett ^77^, Kate F Cook ^84^, Christopher Fearn ^84^, Salman Goudarzi ^84^, Ronan A Lyons ^88^, Thomas Williams ^104^, Sam T Haldenby ^107^, Jillian Durham ^116^ and Steven Leonard ^116^

**Metadata curation, and Software and analysis tools:**

Robert M Davies ^116^

**Project administration, and Samples and logistics:**

Rahul Batra ^12^, Beth Blane ^20^, Moira J Spyer ^30, 95, 96^, Perminder Smith ^32, 112^, Mehmet Yavus ^85, 109^, Rachel J Williams ^96^, Adhyana IK Mahanama ^97^, Buddhini Samaraweera ^97^, Sophia T Girgis ^102^, Samantha E Hansford ^109^, Angie Green ^115^, Charlotte Beaver ^116^, Katherine L Bellis ^116, 102^, Matthew J Dorman ^116^, Sally Kay ^116^, Liam Prestwood ^116^ and Shavanthi Rajatileka ^116^

**Project administration, and Sequencing and analysis:**

Joshua Quick ^43^

**Project administration, and Software and analysis tools:**

Radoslaw Poplawski ^43^

**Samples and logistics, and Sequencing and analysis:**

Nicola Reynolds ^8^, Andrew Mack ^11^, Arthur Morriss ^11^, Thomas Whalley ^11^, Bindi Patel ^12^, Iliana Georgana ^24^, Myra Hosmillo ^24^, Malte L Pinckert ^24^, Joanne Stockton ^43^, John H Henderson ^65^, Amy Hollis ^65^, William Stanley ^65^, Wen C Yew ^65^, Richard Myers ^72^, Alicia Thornton ^72^, Alexander Adams ^74^, Tara Annett ^74^, Hibo Asad ^74^, Alec Birchley ^74^, Jason Coombes ^74^, Johnathan M Evans ^74^, Laia Fina ^74^, Bree Gatica-Wilcox ^74^, Lauren Gilbert ^74^, Lee Graham ^74^, Jessica Hey ^74^, Ember Hilvers ^74^, Sophie Jones ^74^, Hannah Jones ^74^, Sara Kumziene-Summerhayes ^74^, Caoimhe McKerr ^74^, Jessica Powell ^74^, Georgia Pugh ^74^, Sarah Taylor ^74^, Alexander J Trotter ^75^, Charlotte A Williams ^96^, Leanne M Kermack ^102^, Benjamin H Foulkes ^109^, Marta Gallis ^109^, Hailey R Hornsby ^109^, Stavroula F Louka ^109^, Manoj Pohare ^109^, Paige Wolverson ^109^, Peijun Zhang ^109^, George MacIntyre-Cockett ^115^, Amy Trebes ^115^, Robin J Moll ^116^, Lynne Ferguson ^117^, Emily J Goldstein ^117^, Alasdair Maclean ^117^ and Rachael Tomb ^117^

**Samples and logistics, and Software and analysis tools:**

Igor Starinskij ^53^

**Sequencing and analysis, and Software and analysis tools:**

Laura Thomson ^5^, Joel Southgate ^11, 74^, Moritz UG Kraemer ^23^, Jayna Raghwani ^23^, Alex E Zarebski ^23^, Olivia Boyd ^39^, Lily Geidelberg ^39^, Chris J Illingworth ^52^, Chris Jackson ^52^, David Pascall ^52^, Sreenu Vattipally ^53^, Timothy M Freeman ^109^, Sharon N Hsu ^109^, Benjamin B Lindsey ^109^, Keith James ^116^, Kevin Lewis ^116^, Gerry Tonkin-Hill ^116^ and Jaime M Tovar-Corona ^116^

**Sequencing and analysis, and Visualisation:**

MacGregor Cox ^20^

**Software and analysis tools, and Visualisation:**

Khalil Abudahab ^14, 116^, Mirko Menegazzo ^14^, Ben EW Taylor MEng ^14, 116^, Corin A Yeats ^14^, Afrida Mukaddas ^53^, Derek W Wright ^53^, Leonardo de Oliveira Martins ^75^, Rachel Colquhoun ^104^, Verity Hill

^104^, Ben Jackson ^104^, JT McCrone ^104^, Nathan Medd ^104^, Emily Scher ^104^ and Jon-Paul Keatley ^116^

**Leadership and supervision:**

Tanya Curran ^3^, Sian Morgan ^10^, Patrick Maxwell ^20^, Ken Smith ^20^, Sahar Eldirdiri ^21^, Anita Kenyon ^21^, Alison H Holmes ^38, 57^, James R Price ^38, 57^, Tim Wyatt ^69^, Alison E Mather ^75^, Timofey Skvortsov ^77^ and John A Hartley ^96^

**Metadata curation:**

Martyn Guest ^11^, Christine Kitchen ^11^, Ian Merrick ^11^, Robert Munn ^11^, Beatrice Bertolusso ^33^, Jessica Lynch ^33^, Gabrielle Vernet ^33^, Stuart Kirk ^34^, Elizabeth Wastnedge ^56^, Rachael Stanley ^58^, Giles Idle ^64^,Declan T Bradley ^69, 77^, Jennifer Poyner ^79^ and Matilde Mori ^110^

**Project administration:**

Owen Jones ^11^, Victoria Wright ^18^, Ellena Brooks ^20^, Carol M Churcher ^20^, Mireille Fragakis ^20^, Katerina Galai ^20, 70^, Andrew Jermy ^20^, Sarah Judges ^20^, Georgina M McManus ^20^, Kim S Smith ^20^, Elaine Westwick ^20^, Stephen W Attwood ^23^, Frances Bolt ^38, 57^, Alisha Davies ^74^, Elen De Lacy ^74^, Fatima Downing ^74^, Sue Edwards ^74^, Lizzie Meadows ^75^, Sarah Jeremiah ^97^, Nikki Smith ^109^ and Luke Foulser ^116^

**Samples and logistics:**

Themoula Charalampous ^12, 46^, Amita Patel ^12^, Louise Berry ^15^, Tim Boswell ^15^, Vicki M Fleming ^15^, Hannah C Howson-Wells ^15^, Amelia Joseph ^15^, Manjinder Khakh ^15^, Michelle M Lister ^15^, Paul W Bird ^16^, Karlie Fallon ^16^, Thomas Helmer ^16^, Claire L McMurray ^16^, Mina Odedra ^16^, Jessica Shaw ^16^, Julian W Tang ^16^, Nicholas J Willford ^16^, Victoria Blakey ^17^, Veena Raviprakash ^17^, Nicola Sheriff ^17^, Lesley-Anne Williams ^17^, Theresa Feltwell ^20^, Luke Bedford ^26^, James S Cargill ^27^, Warwick Hughes ^27^, Jonathan Moore ^28^, Susanne Stonehouse ^28^, Laura Atkinson ^29^, Jack CD Lee ^29^, Dr Divya Shah ^29^, Adela Alcolea-Medina ^32, 112^, Natasha Ohemeng-Kumi ^32, 112^, John Ramble ^32, 112^, Jasveen Sehmi ^32, 112^, Rebecca Williams ^33^, Wendy Chatterton ^34^, Monika Pusok ^34^, William Everson ^37^, Anibolina Castigador ^44^, Emily Macnaughton ^44^, Kate El Bouzidi ^45^, Temi Lampejo ^45^, Malur Sudhanva ^45^, Cassie Breen ^47^, Graciela Sluga ^48^, Shazaad SY Ahmad ^49, 70^, Ryan P George ^49^, Nicholas W Machin ^49, 70^, Debbie Binns ^50^, Victoria James ^50^, Rachel Blacow ^55^, Lindsay Coupland ^58^, Louise Smith ^59^, Edward Barton ^60^, Debra Padgett ^60^, Garren Scott ^60^, Aidan Cross ^61^, Mariyam Mirfenderesky ^61^, Jane Greenaway ^62^, Kevin Cole ^64^, Phillip Clarke ^67^, Nichola Duckworth ^67^, Sarah Walsh ^67^, Kelly Bicknell ^68^, Robert Impey ^68^, Sarah Wyllie ^68^, Richard Hopes ^70^, Chloe Bishop ^72^, Vicki Chalker ^72^, Ian Harrison ^72^, Laura Gifford ^74^, Zoltan Molnar ^77^, Cressida Auckland ^79^, Cariad Evans ^85, 109^, Kate Johnson ^85, 109^, David G Partridge ^85, 109^, Mohammad Raza ^85, 109^, Paul Baker ^86^, Stephen Bonner ^86^, Sarah Essex ^86^, Leanne J Murray ^86^, Andrew I Lawton ^87^, Shirelle Burton-Fanning ^89^, Brendan AI Payne ^89^, Sheila Waugh ^89^, Andrea N Gomes ^91^, Maimuna Kimuli ^91^, Darren R Murray ^91^, Paula Ashfield ^92^, Donald Dobie ^92^, Fiona Ashford ^93^, Angus Best ^93^, Liam Crawford ^93^, Nicola Cumley ^93^, Megan Mayhew ^93^, Oliver Megram ^93^, Jeremy Mirza ^93^, Emma Moles-Garcia ^93^, Benita Percival ^93^, Megan Driscoll ^96^, Leah Ensell ^96^, Helen L Lowe ^96^, Laurentiu Maftei ^96^, Matteo Mondani ^96^, Nicola J Chaloner ^99^, Benjamin J Cogger ^99^, Lisa J Easton ^99^, Hannah Huckson ^99^, Jonathan Lewis ^99^, Sarah Lowdon ^99^, Cassandra S Malone ^99^, Florence Munemo ^99^, Manasa Mutingwende ^99^, Roberto Nicodemi ^99^, Olga Podplomyk ^99^, Thomas Somassa ^99^, Andrew Beggs ^100^, Alex Richter ^100^, Claire Cormie ^102^, Joana Dias ^102^, Sally Forrest ^102^, Ellen E Higginson ^102^, Mailis Maes ^102^, Jamie Young ^102^, Rose K Davidson ^103^, Kathryn A Jackson ^107^, Lance Turtle ^107^, Alexander J Keeley ^109^, Jonathan Ball ^113^, Timothy Byaruhanga ^113^, Joseph G Chappell ^113^, Jayasree Dey ^113^, Jack D Hill ^113^, Emily J Park ^113^, Arezou Fanaie ^114^, Rachel A Hilson ^114^, Geraldine Yaze ^114^ and Stephanie Lo ^116^

**Sequencing and analysis:**

Safiah Afifi ^10^, Robert Beer ^10^, Joshua Maksimovic ^10^, Kathryn McCluggage ^10^, Karla Spellman ^10^, Catherine Bresner ^11^, William Fuller ^11^, Angela Marchbank ^11^, Trudy Workman ^11^, Ekaterina Shelest ^13,81^, Johnny Debebe ^18^, Fei Sang ^18^, Marina Escalera Zamudio ^23^, Sarah Francois ^23^, Bernardo Gutierrez ^23^, Tetyana I Vasylyeva ^23^, Flavia Flaviani ^31^, Manon Ragonnet-Cronin ^39^, Katherine L Smollett ^42^, Alice Broos ^53^, Daniel Mair ^53^, Jenna Nichols ^53^, Kyriaki Nomikou ^53^, Lily Tong ^53^, Ioulia Tsatsani ^53^, Sarah O’Brien ^54^, Steven Rushton ^54^, Roy Sanderson ^54^, Jon Perkins ^55^, Seb Cotton ^56^, Abbie Gallagher ^56^, Elias Allara ^70, 102^, Clare Pearson ^70, 102^, David Bibby ^72^, Gavin Dabrera ^72^, Nicholas Ellaby ^72^, Eileen Gallagher ^72^, Jonathan Hubb ^72^, Angie Lackenby ^72^, David Lee ^72^, Nikos Manesis ^72^, Tamyo Mbisa ^72^, Steven Platt ^72^, Katherine A Twohig ^72^, Mari Morgan ^74^, Alp Aydin ^75^, David J Baker ^75^, Ebenezer Foster-Nyarko ^75^, Sophie J Prosolek ^75^, Steven Rudder ^75^, Chris Baxter ^77^, Sílvia F Carvalho ^77^, Deborah Lavin ^77^, Arun Mariappan ^77^, Clara Radulescu ^77^, Aditi Singh ^77^, Miao Tang ^77^, Helen Morcrette ^79^, Nadua Bayzid ^96^, Marius Cotic ^96^, Carlos E Balcazar ^104^, Michael D Gallagher ^104^, Daniel Maloney ^104^, Thomas D Stanton ^104^, Kathleen A Williamson ^104^, Robin Manley ^105^, Michelle L Michelsen ^105^, Christine M Sambles ^105^, David J Studholme ^105^, Joanna Warwick-Dugdale ^105^, Richard Eccles ^107^, Matthew Gemmell ^107^, Richard Gregory ^107^, Margaret Hughes ^107^, Charlotte Nelson ^107^, Lucille Rainbow ^107^, Edith E Vamos ^107^, Hermione J Webster ^107^, Mark Whitehead ^107^, Claudia Wierzbicki ^107^, Adrienn Angyal ^109^, Luke R Green ^109^, Max Whiteley ^109^, Emma Betteridge ^116^, Iraad F Bronner ^116^, Ben W Farr ^116^, Scott Goodwin ^116^, Stefanie V Lensing ^116^, Shane A McCarthy ^116, 102^, Michael A Quail ^116^, Diana Rajan ^116^, Nicholas M Redshaw ^116^, Carol Scott ^116^, Lesley Shirley ^116^ and Scott AJ Thurston ^116^

**Software and analysis tools:**

Will Rowe ^43^, Amy Gaskin ^74^, Thanh Le-Viet ^75^, James Bonfield ^116^, Jennifier Liddle ^116^ and Andrew Whitwham ^116^

**1** Barking, Havering and Redbridge University Hospitals NHS Trust, **2** Barts Health NHS Trust, **3** Belfast Health & Social Care Trust, **4** Betsi Cadwaladr University Health Board, **5** Big Data Institute, Nuffield Department of Medicine, University of Oxford, **6** Blackpool Teaching Hospitals NHS Foundation Trust, **7** Bournemouth University, **8** Cambridge Stem Cell Institute, University of Cambridge, **9** Cambridge University Hospitals NHS Foundation Trust, **10** Cardiff and Vale University Health Board, **11** Cardiff University, **12** Centre for Clinical Infection and Diagnostics Research, Department of Infectious Diseases, Guy’s and St Thomas’ NHS Foundation Trust, **13** Centre for Enzyme Innovation, University of Portsmouth, **14** Centre for Genomic Pathogen Surveillance, University of Oxford, **15** Clinical Microbiology Department, Queens Medical Centre, Nottingham University Hospitals NHS Trust, **16** Clinical Microbiology, University Hospitals of Leicester NHS Trust, **17** County Durham and Darlington NHS Foundation Trust, **18** Deep Seq, School of Life Sciences, Queens Medical Centre, University of Nottingham, **19** Department of Infectious Diseases and Microbiology, Cambridge University Hospitals NHS Foundation Trust, **20** Department of Medicine, University of Cambridge, **21** Department of Microbiology, Kettering General Hospital, **22** Department of Microbiology, South West London Pathology, **23** Department of Zoology, University of Oxford, **24** Division of Virology, Department of Pathology, University of Cambridge, **25** East Kent Hospitals University NHS Foundation Trust, **26** East Suffolk and North Essex NHS Foundation Trust, **27** East Sussex Healthcare NHS Trust, **28** Gateshead Health NHS Foundation Trust, **29** Great Ormond Street Hospital for Children NHS Foundation Trust, **30** Great Ormond Street Institute of Child Health (GOS ICH), University College London (UCL), **31** Guy’s and St. Thomas’ Biomedical Research Centre, **32** Guy’s and St. Thomas’ NHS Foundation Trust, **33** Hampshire Hospitals NHS Foundation Trust, **34** Health Services Laboratories, **35** Heartlands Hospital, Birmingham, **36** Hub for Biotechnology in the Built Environment, Northumbria University, **37** Hull University Teaching Hospitals NHS Trust, **38** Imperial College Healthcare NHS Trust, **39** Imperial College London, **40** Infection Care Group, St George’s University Hospitals NHS Foundation Trust, **41** Institute for Infection and Immunity, St George’s University of London, **42** Institute of Biodiversity, Animal Health & Comparative Medicine, **43** Institute of Microbiology and Infection, University of Birmingham, **44** Isle of Wight NHS Trust, **45** King’s College Hospital NHS Foundation Trust, **46** King’s College London, **47** Liverpool Clinical Laboratories, **48** Maidstone and Tunbridge Wells NHS Trust, **49** Manchester University NHS Foundation Trust, **50** Microbiology Department, Buckinghamshire Healthcare NHS Trust, **51** Microbiology, Royal Oldham Hospital, **52** MRC Biostatistics Unit, University of Cambridge, **53** MRC-University of Glasgow Centre for Virus Research, **54** Newcastle University, **55** NHS Greater Glasgow and Clyde, **56** NHS Lothian, **57** NIHR Health Protection Research Unit in HCAI and AMR, Imperial College London, **58** Norfolk and Norwich University Hospitals NHS Foundation Trust, **59** Norfolk County Council, **60** North Cumbria Integrated Care NHS Foundation Trust, **61** North Middlesex University Hospital NHS Trust, **62** North Tees and Hartlepool NHS Foundation Trust, **63** North West London Pathology, **64** Northumbria Healthcare NHS Foundation Trust, **65** Northumbria University, **66** NU-OMICS, Northumbria University, **67** Path Links, Northern Lincolnshire and Goole NHS Foundation Trust, **68** Portsmouth Hospitals University NHS Trust, **69** Public Health Agency, Northern Ireland, **70** Public Health England, **71** Public Health England, Cambridge, **72** Public Health England, Colindale, **73** Public Health Scotland, **74** Public Health Wales, **75** Quadram Institute Bioscience, **76** Queen Elizabeth Hospital, Birmingham, **77** Queen’s University Belfast, **78** Royal Brompton and Harefield Hospitals, **79** Royal Devon and Exeter NHS Foundation Trust, **80** Royal Free London NHS Foundation Trust, **81** School of Biological Sciences, University of Portsmouth, **82** School of Health Sciences, University of Southampton, **83** School of Medicine, University of Southampton, **84** School of Pharmacy & Biomedical Sciences, University of Portsmouth, **85** Sheffield Teaching Hospitals NHS Foundation Trust, **86** South Tees Hospitals NHS Foundation Trust, **87** Southwest Pathology Services, **88** Swansea University, **89** The Newcastle upon Tyne Hospitals NHS Foundation Trust, **90** The Queen Elizabeth Hospital King’s Lynn NHS Foundation Trust, **91** The Royal Marsden NHS Foundation Trust, **92** The Royal Wolverhampton NHS Trust, **93** Turnkey Laboratory, University of Birmingham, **94** University College London Division of Infection and Immunity, **95** University College London Hospital Advanced Pathogen Diagnostics Unit, **96** University College London Hospitals NHS Foundation Trust, **97** University Hospital Southampton NHS Foundation Trust, **98** University Hospitals Dorset NHS Foundation Trust, **99** University Hospitals Sussex NHS Foundation Trust, **100** University of Birmingham, **101** University of Brighton, **102** University of Cambridge, **103** University of East Anglia, **104** University of Edinburgh, **105** University of Exeter, **106** University of Kent, **107** University of Liverpool, **108** University of Oxford, **109** University of Sheffield, **110** University of Southampton, **111** University of St Andrews, **112** Viapath, Guy’s and St Thomas’ NHS Foundation Trust, and King’s College Hospital NHS Foundation Trust, **113** Virology, School of Life Sciences, Queens Medical Centre, University of Nottingham, **114** Watford General Hospital, **115** Wellcome Centre for Human Genetics, Nuffield Department of Medicine, University of Oxford, **116** Wellcome Sanger Institute, **117** West of Scotland Specialist Virology Centre, NHS Greater Glasgow and Clyde, **118** Whittington Health NHS Trust

## Acknowledgements

COG-UK is supported by funding from the Medical Research Council (MRC) part of UK Research & Innovation (UKRI), the National Institute of Health Research (NIHR) [grant code: MC_PC_19027], and Genome Research Limited, operating as the Wellcome Sanger Institute.

## Data Availability Statement

The data that support the findings of this study are available at Wellcome Sanger Institute COVID–19 Genomic Surveillance (https://covid19.sanger.ac.uk/lineages/raw).

